# Electronic health record decision support for the diagnosis and management of pediatric tuberculosis infection

**DOI:** 10.64898/2026.02.09.26345927

**Authors:** Naythra Narayanan, Matthew T. Murrill, William Burrough, Tessa Mochizuki, Colleen Panina, Mariamawit Tamerat, Julia Fink, Iris L. Wu, Katya Salcedo, Shereen S. Katrak, Terry Mayo, Amit Chitnis, Charlotte Hsieh, Zarin Noor, Gena Lewis, Devan Jaganath

## Abstract

**Objective:** To evaluate whether new tuberculosis (TB) tools in the electronic health record (EHR) can support latent TB infection (LTBI) screening, testing and treatment among children and adolescents in a primary care setting.

**Study Design:** This retrospective cohort study included children and adolescents between the ages 1-25 years who had a well-child or well-adolescent visit at a Federally Qualified Health Center in Oakland, California, from December 2021 to December 2022. Four new EHR tools were introduced for the completion and documentation of TB risk factor screening, testing and treatment. Data were extracted from the EHR to identify gaps in these steps, and logistic regression was used to examine factors associated with completion of TB infection screening and testing. Acceptability was evaluated using provider satisfaction surveys before and after the implementation of TB EHR tools.

**Results:** Of 5,879 individuals (median age of 9 years at first visit, interquartile range (IQR) 4-13 years), 94% completed TB risk factor screening. Among those with a new risk factor, 59% had a TB infection test ordered and 96% completed testing. Ten participants (3%) tested positive, all initiated LTBI treatment, and most (n=7, 70%) completed treatment. Overall, 5,162 (88%) individuals completed their LTBI care cascade. Younger children ages 1-4 years were more likely to be screened for TB risk factors, but were less likely to be tested. Provider satisfaction increased from 40% to 71% for risk factor screening, and 36% to 77% for test ordering.

**Conclusion:** EHR tools supported completion of the pediatric LTBI care cascade, while also increasing provider satisfaction. EHR-based solutions show promise as part of multi-component strategies to address gaps in LTBI care for children and adolescents.

## INTRODUCTION

Tuberculosis (TB) disease is the leading cause of death by an infectious disease globally, and an estimated quarter of the world’s population has TB infection (also known as latent TB infection or LTBI).^1^ As the majority of TB disease (∼80%) in the United States (US) is attributed to the reactivation of asymptomatic TB infection, the screening and treatment of LTBI is critical for TB disease elimination in the US.^2^ California has the largest number of TB disease diagnoses of all US states,^3^ and an estimated more than two million Californians have LTBI.^3^ Because children have many remaining years at risk for TB reactivation and a higher likelihood of severe disease early in life,^4,5^ the lifetime benefit of LTBI treatment is substantial and treatment is typically better tolerated than in adults. Consequently, the American Academy of Pediatrics (AAP) recommends annual TB risk factor screening at well-child and well-adolescent visits, testing those at increased risk of TB with an interferon gamma release assay (IGRA) or tuberculin skin test (TST), and initiating LTBI treatment for those with positive testing and no evidence of TB disease.^6^

There is limited evidence, however, on how to effectively implement pediatric TB risk factor screening, infection testing, and LTBI treatment in the US. Recent analyses have identified large gaps in the implementation of TB preventive care, particularly TB infection testing and treatment initiation.^7–10^ We previously examined LTBI care at a pediatric Federally Qualified Health Center (FQHC) in Northern California from 2014-2020, and showed that only 20% of children with TB risk factors were tested with an IGRA or TST.^9^ At the same time, we found that adding TB risk factor questions to a well-child and well-adolescent note template can significantly increase risk factor screening.^9^

Consequently, we created and implemented new LTBI EHR tools within Epic (Epic Systems, Verona, Wisconsin) for TB risk factor screening, clinical decision support and documentation. In this study, we evaluated TB preventive care from 2021-2022 after introduction of this workflow to examine whether these tools could support monitoring and completion of LTBI care, increase provider satisfaction with clinical workflows, and identify ongoing needs to improve the quality of TB preventive care.

## METHODS

### Study Setting

The pediatric FQHC is located in Oakland, California.^11^ Oakland is an urban city located in Alameda County, which had a TB disease incidence of 7.8 diagnoses per 100,000 residents in 2024, the fourth highest among all California counties. Approximately 9% of TB disease diagnoses in Alameda County in 2024 also occurred among individuals under 24 years of age.^12^ The FQHC is part of University of California San Francisco (UCSF) Benioff Children’s Hospitals and UCSF Health, which uses the Epic EHR platform.

### TB Infection Clinical Support Tools

The FQHC had previously introduced TB risk factor questions into the well-child or well-adolescent progress note templates, starting in 2014. In 2021, we met with clinic leadership and providers and received feedback on approaches to expand TB clinical decision support within the EHR. We then created mock-ups of their ideas, and presented them at follow-up meetings for iterative feedback. After these were created in the EHR, we then shared the workflow with clinic providers for final feedback before placing them in production in December 2021.

The new EHR tools were (**Supplemental File 1**):

1. **TB Risk Factor Screening Care Gap**. Care gaps list pending health maintenance tasks in the patient chart. They are located on a patient’s storyboard (i.e., chart summary), a panel of the EHR that is visible whenever a chart is opened, as well as under the health maintenance tab. We created a new TB risk factor screening care gap that appeared annually, which when clicked displayed past risk factor questionnaires and provided links to institutional and public health guidelines. The care gap could be completed if a TB risk factor questionnaire was completed or if manually postponed with a reason indicated.
2. **TB Risk Factor Questionnaire**. Four TB risk factor questions, aligned with the California pediatric TB risk assessment were located in the “Rooming” tab for the visit: 1) birth outside of the US in a TB endemic country (i.e., countries other than Canada, Western or Northern Europe, Australia or New Zealand; 2) travel or residence for ≥1 month in a TB endemic country; 3) current or planned immunosuppression; or 4) known close contact with someone with TB disease.^13,14^ These questions could be completed by providers or other clinic staff.
3. **Best Practice Advisory (BPA) for TB Infection Testing**. If any of the above risk factors were selected, a BPA appeared in the “Plan” tab to notify the provider that a test for TB infection was indicated. The BPA included options to order a blood-based QuantiFERON Gold-in-Tube (QFT) or tuberculin skin test (TST for children under 2 years), or a provider could acknowledge the BPA and provide a reason for not ordering a test.
4. **Specialty Navigator**. A TB specialty navigator was created so that providers could document TB infection and chest X-ray (CXR) results, LTBI treatment decisions, and completion of treatment. This was created to allow tracking and communication of LTBI care between providers and for easier reference in the future, and this information could be automatically brought into the progress note.

Prior to implementation, we gave a presentation on TB care and an overview of these new tools to all clinic providers and provided the clinic an instructions document to guide use. Clinic leadership who helped to develop the tools were also available to answer any questions and/or provide assistance.

### Adoption and Acceptability

To examine adoption, we calculated the proportion of visits in which each EHR tool was used, overall and the monthly change. To assess provider acceptability, an anonymous provider survey was administered before and six months after the implementation of the LTBI EHR tools to assess satisfaction **(Supplemental File 2**). Questions used a 5-point Likert scale ranging from extremely satisfied to extremely dissatisfied. For our analysis, we defined satisfied as extremely or somewhat satisfied. We summarized the proportion of satisfaction with each aspect of LTBI evaluation and management, before and after implementation of the EHR tools.

### LTBI Care Cascade

To evaluate gaps in TB preventive care after the implementation of clinical decision support tools in the EHR, we created an LTBI care cascade including all individuals 1-25 years of age who were receiving care at the FQHC and had at least one well-child or well-adolescent visit from December 2021 through December 2022. We excluded individuals if a provider manually postponed their care gap during the intervention period, or with a history of LTBI or TB disease diagnosis prior to December 2021, based on encounter diagnoses, review of visit notes, or previously positive QFT or TST. To determine eligibility, we used extracted EHR data from all clinic visits during this period as well as testing and treatment data prior to visits (From July 2011). We also extracted LTBI treatment and testing data through December 2023 to collect information on any follow-up care after the visits.

### EHR Data Extraction

For all eligible individuals, we obtained sociodemographic data including age, sex, ethnicity, race, preferred language, and insurance. Documentation of TB risk factors was obtained from the TB risk assessment questionnaire, as well as progress notes by extracting any text on TB risk factors. Ordering and results of TB infection testing and CXRs were extracted along with responses to each clinical support tool, and any reasons for not testing. We extracted all TB infection and TB disease medications prescribed during the study period, including isoniazid, rifampin, rifapentine, ethambutol, rifabutin, pyrazinamide, and levofloxacin. Among individuals who were prescribed these medications, chart review was conducted to then confirm their LTBI diagnosis, treatment regimen and outcome of completion.

### Analysis

To construct an LTBI care cascade, we measured the number of individuals completing the following steps: (1) identifying individuals with new TB risk factors through a risk assessment questionnaire, (2) ordering TB infection testing of individuals with new risks (QFT or TST), (3) completing testing, (4) CXR ordering for those with a positive TB infection test to rule out TB disease, (5) LTBI treatment initiation, and (6) LTBI treatment completion. For steps 2 and 3, we assessed provider ordering and patient completion separately. The care cascade was constructed in an incremental manner for all patient visits (**Table 1**). Gaps in the care cascade were defined as the proportion of individuals who were eligible to and did not complete a given step. We constructed the overall care cascade, as well as by age category (1-4 years old, 5-12 years old, and 13+ years old) and primary language (English, Spanish, Other). Univariate logistic regression models were constructed to assess factors associated with completion of the TB risk assessment and TB testing. The odds ratios (ORs) were reported with a 95% confidence interval (CI) and significance was defined as a p-value < 0.05. Statistical analysis was conducted in R version 4.4.3.

**Table 1.** Definition of TB care cascade steps.

| Numerator Definition | Numerator Total | Denominator Definition | Denominator Total |
| --- | --- | --- | --- |
| Number of eligible individuals | 5,879 | -- | -- |
| Individuals who had a TB risk factor | 699 | Number of eligible individuals | 5,879 |
| Individuals who had a new risk factor.<sup>1</sup> | 626 | The number of individuals who had at least one TB risk factor during the study. | 699 |
| The number of individuals who had a TB test ordered | 369 | The number of individuals who had a new risk factor | 626 |
| The number of individuals who had a TB test result | 355 | The number of individuals who had a TB test ordered | 369 |
| The number of individuals who had a chest X-ray ordered | 10 | The number of individuals who tested positive for TB | 10 |
| The number of individuals who started LTBI treatment | 10 | The number of individuals who had a chest X-ray | 10 |
| The number of individuals who completed LTBI treatment | 7 | The number of individuals who started LTBI treatment | 10 |

- 415 1. Those who visited the clinic and are born outside the US, traveled outside the US for greater than one month, are or planned to be immunosuppressed, or in contact with a person with TB disease are included as having a new risk factor. Individuals whose only risk is being born outside the US and have previously tested negative for TB infection by TST or QFT are not included in the numerator as they do not have a new TB risk factor.

### Ethical Considerations

This study was reviewed and approved by the Institutional Review Board (IRB) of the University of California, San Francisco.

## RESULTS

### Study Population

From December 10, 2021 to December 8, 2022, there were 5,879 unique patients who met the inclusion criteria and presented to the FQHC for a well-child or -adolescent visit (**Figure 1**). Demographic characteristics are shown in **Table 2**. The median age of participants at their first visit was 9 years old, 49% females and 51% were males. Most participants spoke English as their primary language (72%), followed by Spanish (13%). Most families were self-reported Black/African American (39%), and most used public insurance (State Medicaid, 89%).

**Figure 1.**
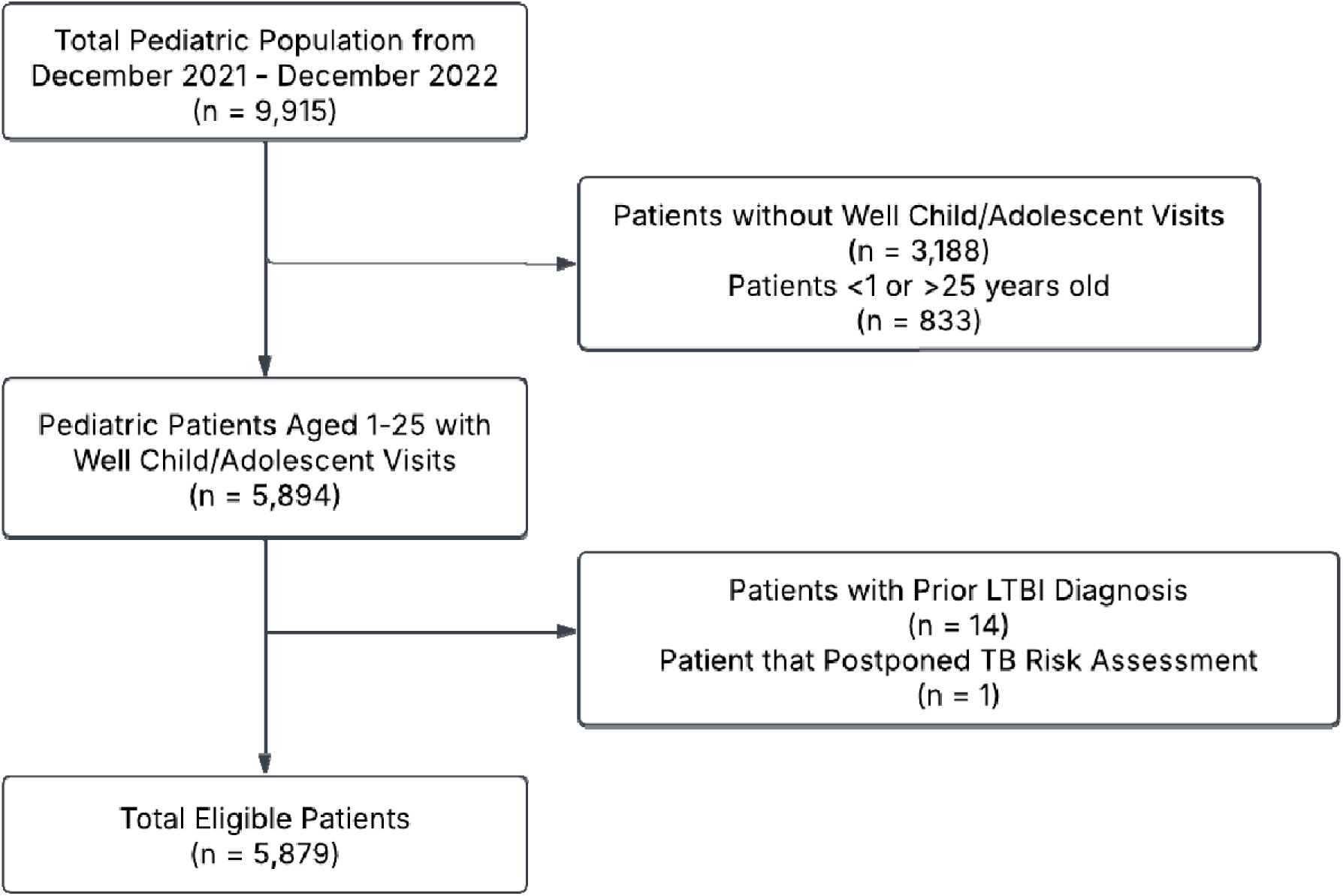
Flowchart of cohort included in analysis

**Table 2.** Cohort Demographics.

| Characteristics | Patients (N=5,879) |
| --- | --- |
| Age at first visit, median years (IQR) | 9 (4-13) |
| Age category at first visit n (%) |  |
| 1-4 years | 1,552 (26.4) |
| 5-10 years | 2,028 (34.5) |
| 11-15 years | 1,523 (25.9) |
| 16-20 years | 748 (12.7) |
| 20-25 years | 28 (0.5) |
| Gender n (%) |  |
| Female | 2,854 (48.5) |
| Male | 3,004 (51.1) |
| Nonbinary | 1 (0.02) |
| Not Reported | 20 (0.3) |
| Race/Ethnicity n (%) |  |
| Asian | 618 (10.5) |
| Black/African American | 2,262 (38.5) |
| Hispanic | 1,757 (29.9) |
| Native American/Alaskan Native | 3 (0.05) |
| Native Hawaiian/Other Pacific Islander | 34 (0.6) |
| White | 289 (4.9) |
| Multi-Race/Ethnicity | 218 (3.7) |
| Other/Unknown | 698 (11.9) |
| Primary Language n (%) |  |
| English | 4,253 (72.3) |
| Spanish | 781 (13.3) |
| Arabic | 180 (3.1) |
| Tigrinya | 110 (1.9) |
| Other | 224 (3.8) |
| Missing/Declined | 331 (5.6) |
| Insurance n (%) |  |
| Public | 5,202 (88.5) |
| Private | 661 (11.2) |
| No Insurance | 16 (0.3) |

IQR – Interquartile Range

### LTBI EHR Tool Adoption

Adoption of the EHR tools is shown in **Figure 2**. Most providers (94%) used either the new TB questionnaire or the existing progress note template to complete TB risk factor screening in similar proportions (**Figure 2A**), and this remained stable over time (**Figure 2B**). Among those with a new TB risk factor, 28% used the BPA to order a test or acknowledge a reason (**Figure 2C**). An additional 40% ordered a test outside of the BPA, and 32% neither used the BPA nor ordered a test. Lastly, among the ten children with a positive test result, six (60%) had data entered in the TB specialty navigator, and three of the four without data were lost to follow up or moved out of the area. Four of the six (67%) had documentation of treatment completion in the navigator.

**Figure 2.**
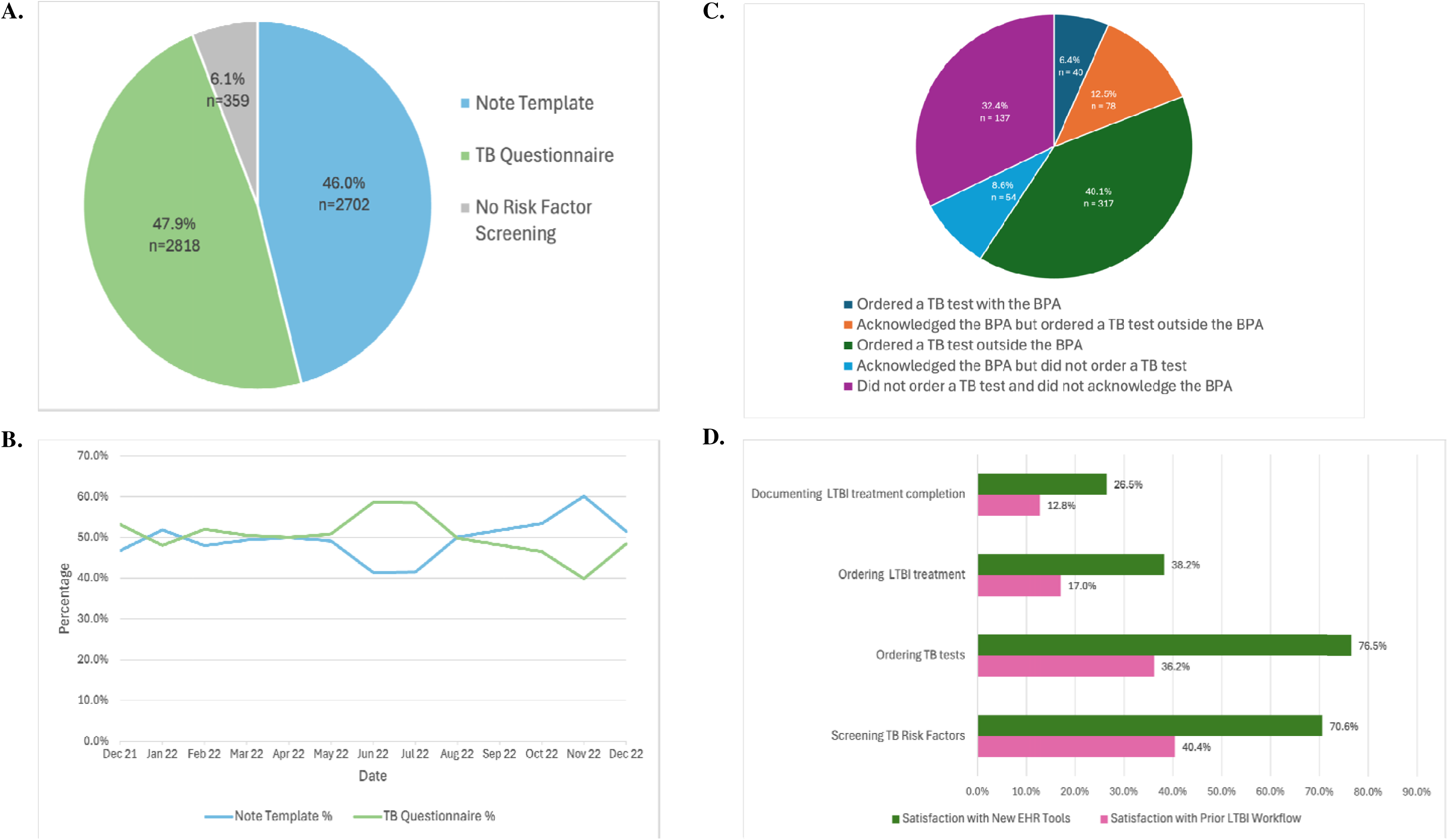
Adoption and Acceptability of Electronic Health Record tools for TB. A) Overall usage of a note template versus a TB questionnaire for risk factor screening; B) Note template versus TB questionnaire usage for risk factor screening over time; C) Usage of the best practice advisory for TB infection test ordering; D) Provider satisfaction of LTBI workflow, pre- and post-implementation of the new electronic health record tools.

### LTBI EHR Tool Acceptability

Of 118 providers, 47 (40%) answered the pre-implementation survey and 34 (29%) answered the post-implementation survey (**Figure 2D**). Overall, providers were more satisfied with the new EHR tools for screening TB risk factors, ordering TB tests, ordering LTBI treatment, and documenting LTBI treatment completion. Providers were particularly satisfied with the new EHR tools for screening TB risk factors (71%) and ordering TB tests (77%). While increased, satisfaction with documentation of treatment completion and ordering LTBI treatment remained low at 27% and 38%, respectively.

### TB Infection Care Cascade

The LTBI care cascade is shown in **Figure 3**. Overall, 5,520 of eligible 5,879 patients (94%) completed the TB risk assessment and 699 (13%) reported a risk factor. The most commonly reported risk factors were birth outside the US (285/699, 41%) and travel outside the US for at least one month (214/699, 31%). Of the participants that screened positive for the risk assessment, 73 individuals (10%) did not have a new TB risk factor as their only TB risk was birth outside the US, and they had a previously documented negative QFT or TST result. Of the remaining 626, 369 (59%) children and adolescents had a TB test ordered and 355 (96%) completed the TB test. Nearly all were tested with a QFT (352/355, 99%). Among the participants that had a completed TB infection test, 10 individuals tested positive (3%), and they all received chest X-rays and were started on four months of rifampin to treat LTBI. Seven of the 10 children (70%) completed treatment, and the other three were lost to follow up. In total, 5,162 of 5,879 (88%) of eligible individuals completed their LTBI care cascade.

**Figure 3.**
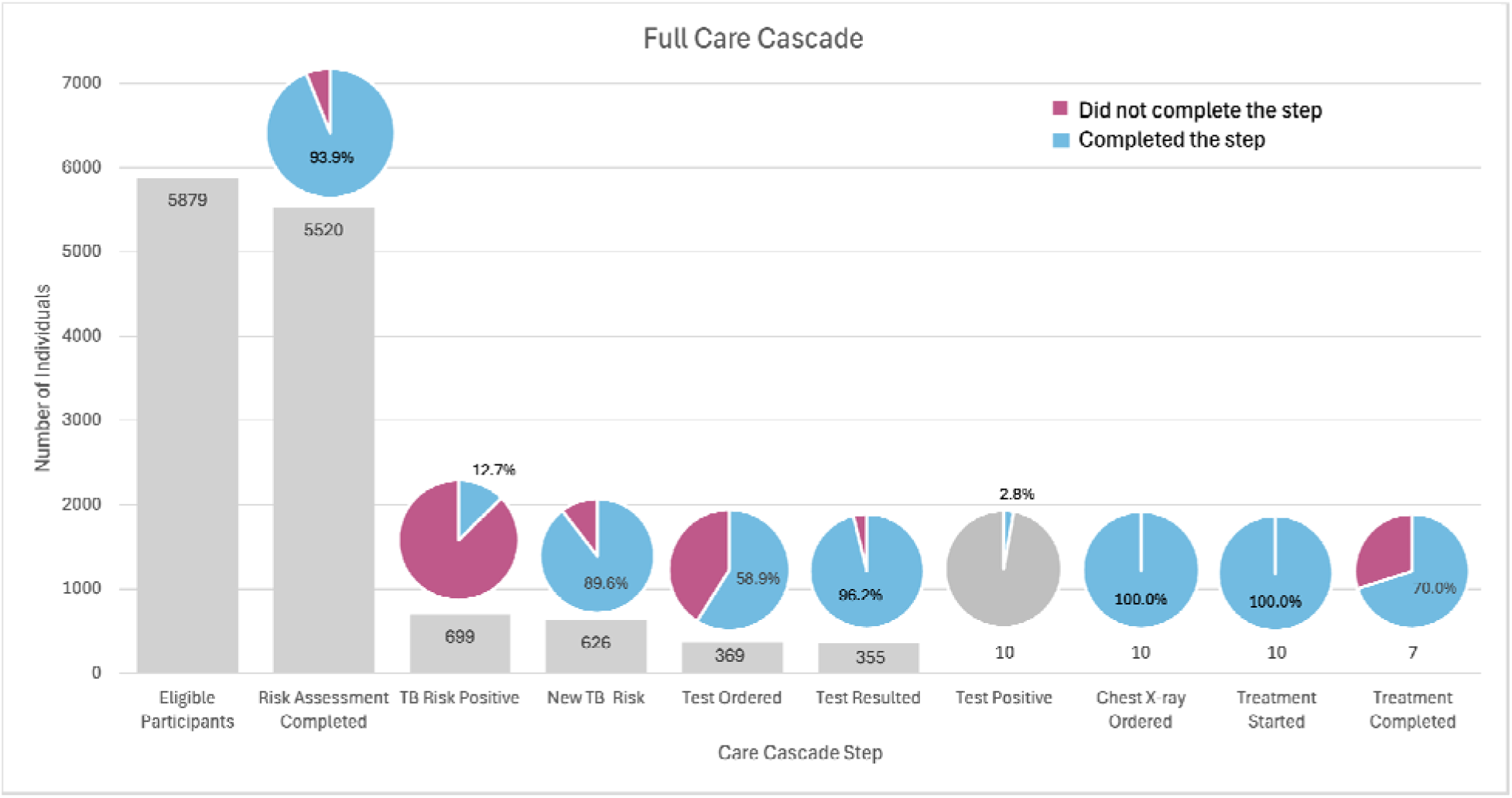
Care Cascade for pediatric LTBI (n = 5,879 participants). Each bar represents the number of individuals where the step was completed, and is the denominator for the next step.

The care cascades by age category (1-4, 5-12, 13+ years) and primary language (English versus non-English) are found in **Supplemental Figures 1** and 2. The care cascades were similar, with high risk factor screening and about half with identified risk factors tested for TB infection.

Univariate logistic regression analyses of the demographic variables associated with TB risk factor screening and TB infection testing are shown in **Table 3**. Individuals aged 1-4 years were more likely to be screened for TB risk factors compared to any other age group, but conversely older age groups were more likely to be tested if a TB risk factor was present. There was no significant difference in completing a TB risk factor questionnaire by race/ethnicity, however those who identified as Asian or Other were more likely to have a TB infection test ordered. Individuals whose primary language was not English were also more likely to be tested for TB infection compared to those whose primary language was English.

**Table 3.**
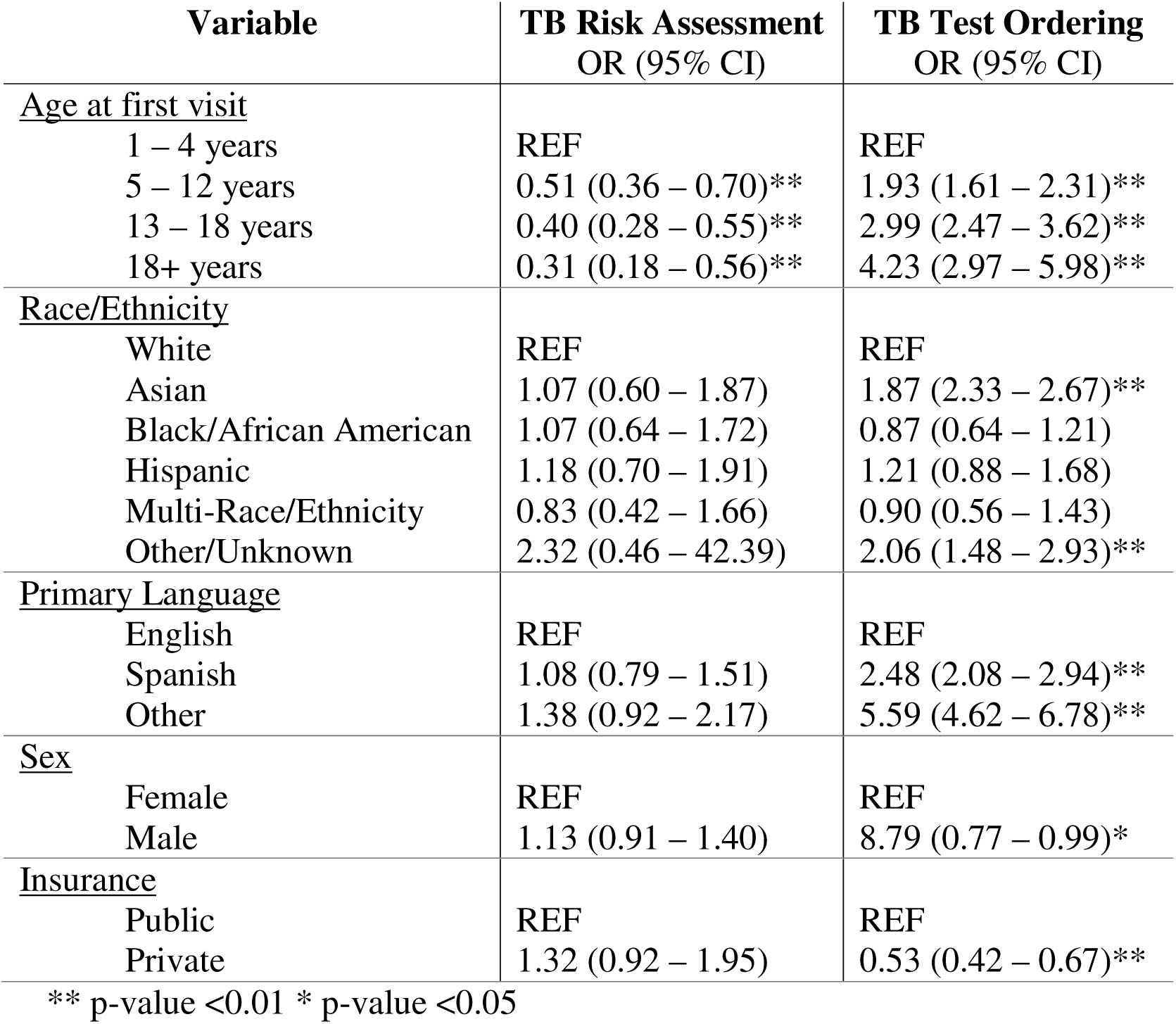
Univariate analysis of factors associated with TB Screening.

## DISCUSSION

While the AAP recommends annual TB risk factor screening for children and adolescents and treatment for those diagnosed with LTBI,^5^ there are significant gaps in the pediatric LTBI care cascade.^7–9^ To address these gaps, we developed and implemented new EHR tools to support primary care providers to deliver TB preventive care. In the year after implementation, provider satisfaction with the TB workflow was higher, in particular for screening and testing. We found that 88% of eligible children and adolescents completed the entire LTBI care cascade, with particularly high risk factor screening completion. Almost 60% with an identified TB risk factor completed TB infection testing, and if positive 70% of children and adolescents completed LTBI treatment. While gaps remain, our results suggest that EHR tools could serve an important role in multi-component interventions to prevent TB disease in children and adolescents.

Provider satisfaction with the TB care workflow was higher after implementation of the new TB EHR tools. Previous studies have shown that provider satisfaction with EHR-based clinical decision support can be variable, depending on provider perception of utility, ease of use, time burden, and alert fatigue.^15–17^ We sought to maximize the benefit of these tools by designing them with clinic leadership and providers with iterative feedback. To assess provider satisfaction, we used an anonymous survey to encourage participation and reduce social desirability bias; however, anonymizing prevented a paired comparison of satisfaction pre- and post-implementation. Notably, provider satisfaction was highest for TB risk factor screening and TB infection, while still low for LTBI treatment initiation and documentation of treatment completion (<50%). This is consistent with the EHR tools that were primarily implemented on risk factor screening and testing. Next steps should focus on challenges in LTBI treatment with ongoing partnership with providers and key stakeholders.

By creating a care gap for every patient in the clinic, all providers had an opportunity to use the new workflow. At the same time, about half of providers adopted the new EHR workflow for TB risk factor screening. Six percent with a risk factor ordered a test within the BPA, but it is possible that it triggered a reminder to order a test and only a third did not order TB infection testing or acknowledge a reason. This reflects the real-world uptake of EHR-based decision support by primary care providers^18,19^ and there will be a range of experience and comfort with LTBI management and EHR tools. Providers that have already established an effective workflow for LTBI care should have the opportunity to continue their approach, while offering additional tools to support providers with less experience to address gaps in TB risk factor screening, testing and treatment.^15^ TB risk factor screening was already high (>90%) prior to the implementation of new EHR tools, and this should be maintained. At the same time, ongoing work is needed to explore any barriers to using these tools, such as provider perceptions of benefit, alert fatigue and computer and EHR literacy.^15^

In a previous retrospective cohort analysis conducted at the same pediatric FQHC from 2014-2020, TB risk assessment completion was high (90% of eligible individuals), but only 20% with a risk factor completed TB infection testing.^9^ With the introduction of new EHR tools, we found that TB risk assessment screening remained high (94%), but TB infection testing increased to nearly 60%. The reasons for this increase are likely multi-factorial. First, the care gap may have created greater awareness around TB risk factor screening and motivated testing. Second, the BPA may have provided a reminder and guidance to order a TB infection test. Third, the provider education on LTBI may have increased knowledge in TB infection evaluation and management. A similar effect was seen at a community health facility in Northern California that predominantly serves a non-US born Asian population.^8^ They found that EHR tools, as part of a multi-component intervention that included provider education, increased TB infection testing in non-US born children from 23% to 80%. Taken together, this suggests that increased training with EHR-based reminders and decision support can be effective to increase TB infection testing in children and adolescents.

In subgroup analyses, younger children 1-4 years old were more likely to be screened for TB risk factors, but were less likely to be tested for TB infection if a risk factor was present. Moreover, non-English speakers were more likely to be tested for TB infection compared with English speakers. These associations were also seen in our previous analyses and in a Boston study which found that adolescents and non-English speakers were more likely to be tested with an IGRA.^13^ There has been limited formative work to explore barriers and facilitators to TB infection testing in children and adolescents, as the majority of studies have focused on steps after testing.^20^ Multi-level barriers likely include provider, caregiver, and patient perceptions of risks and benefits as well as wider challenges in obtaining a blood sample in younger children and costs for testing.

A strength of our study was that we collected data along the full LTBI care cascade at a large FQHC for pediatric primary care in the US. Prior care cascades have started with TB infection testing or were conducted in TB endemic settings.^20^ This is also the first known study to examine new tools for pediatric TB care in the widely-used Epic EHR platform. Limitations of this study include its short time frame and the focus on a single clinic, which could limit generalizability of findings. The response rate for the satisfaction survey was less than 50%, which could have biased the results in favor or against EHR-based TB solutions. These tools are notably now being implemented in other clinics in our health system, and we plan to evaluate their broader impact on longitudinal changes in the LTBI care cascade in the future. This will also facilitate the assessment of later stages in the care cascade (e.g. LTBI treatment completion), as our current study had few individuals with positive test results. Additional challenges with the use of EHR data include missing and incomplete information that can overestimate gaps in the care cascade. To mitigate these potential issues in the present study, particularly for LTBI diagnosis and treatment, we conducted chart review for all individuals with a positive TB infection test.

In conclusion, we found that the implementation of EHR tools for TB prevention showed substantial promise in improving the care cascade for children and adolescents while increasing provider satisfaction in clinic LTBI workflows. Although gaps in TB preventive care for this population persist, these findings support the development and implementation of EHR-based solutions as part of a multi-component strategy to address the barriers to completion of LTBI care in children.

## Data Availability

All data produced in the present study are available upon reasonable request to the authors.

