## Supplemental for "Electronic health record decision support for the diagnosis and management of pediatric tuberculosis infection"

### SUPPLEMENTARY MATERIAL

**Supplemental File 1.** Screenshots of tools created in the electronic health record for tuberculosis infection care.

#### TB Risk Screening Care Gap

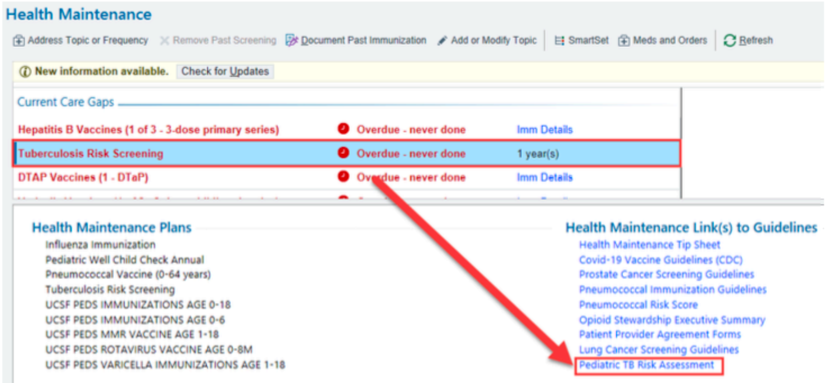


#### TB Screening Questions in the Rooming Tab

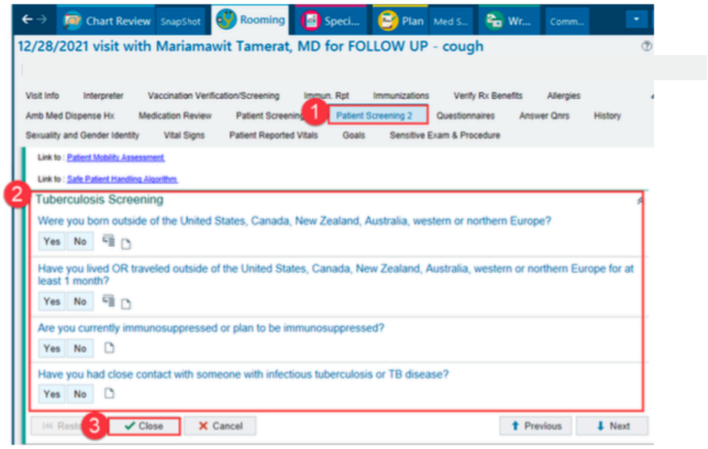


#### Best Practice Advisory

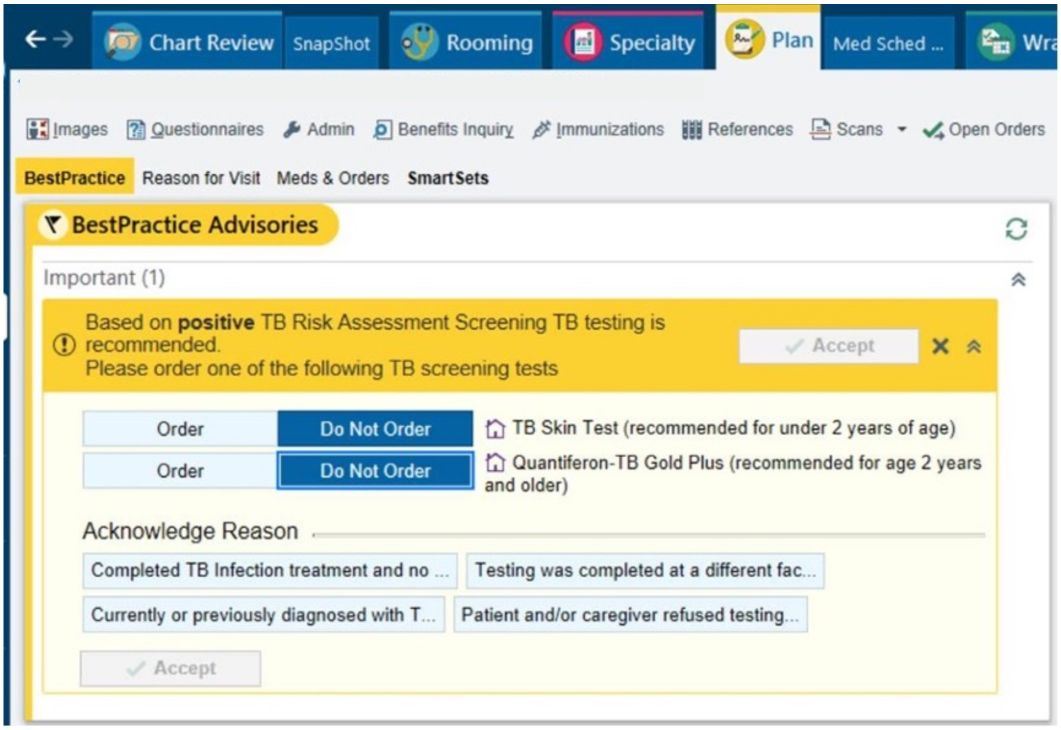


### Specialty Navigator

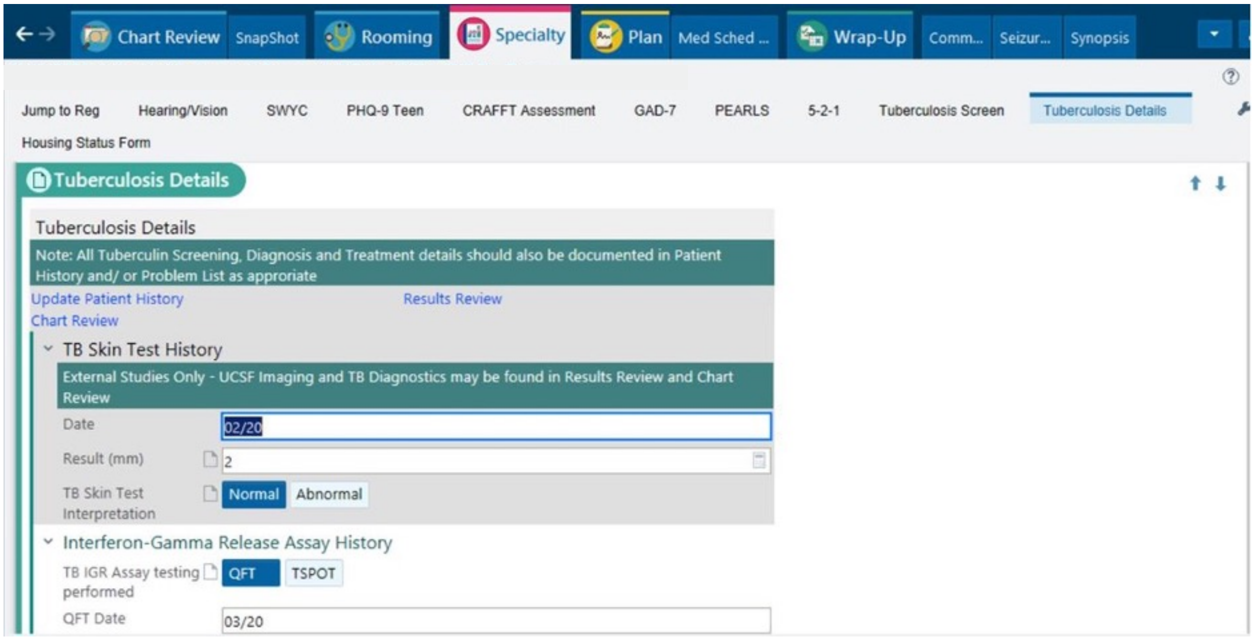

QFT IGR Assay Interp: Positive | **Negative** | Indeterminant

### Imaging

TB Imaging Performed: **Chest X-ray** | Other Imaging

Chest X-ray Date: 04/20

Chest X-ray Result: **Yes** | No

### TB Bacteria Testing

Type: PCR | AFB Smear | AFB Culture

### Diagnosis and Treatment

Diagnosis: **LTBI Diagnosis** | Active Pulmonary TB Diagnosis | Active Extrapulmonary TB Diagnosis

LTBI Treatment Regimen:
- **Daily Rifampin for 4 months (4R)**
- Weekly Isoniazid and Rifapentine for 12 weeks (3HP)
- Daily Isoniazid and Rifampin for 3 months (3HR)
- Isoniazid for 6-9 months (9H)

LTBI Treatment Start Date: 01/19

LTBI Treatment Stopped/ Completion Date: 05/19

LTBI Treatment Completed: **Yes** | No

Close | Cancel | Previous | Next

**Supplemental File 2.** Survey to assess provider satisfaction

How satisfied are you with the following workflows in your electronic medical record (EMR):

| | Very dissatisfied (1) | Somewhat dissatisfied (2) | Neither satisfied nor dissatisfied (3) | Somewhat satisfied (4) | Very satisfied (5) |
|---|---|---|---|---|---|
| Screening of latent TB risk factors (1) | ○ | ○ | ○ | ○ | ○ |
| Ordering TB tests (2) | ○ | ○ | ○ | ○ | ○ |
| Ordering TB treatment (3) | ○ | ○ | ○ | ○ | ○ |
| Documenting latent TB infection treatment completion (4) | ○ | ○ | ○ | ○ | ○ |

**Supplemental Figure 1.** Care Cascade for pediatric LTBI by age group: A) 1 – 4 years old (n = 1,552); B) 5-12 years (n = 2,655); C) ≥13 years (n = 1,672)

**A.**

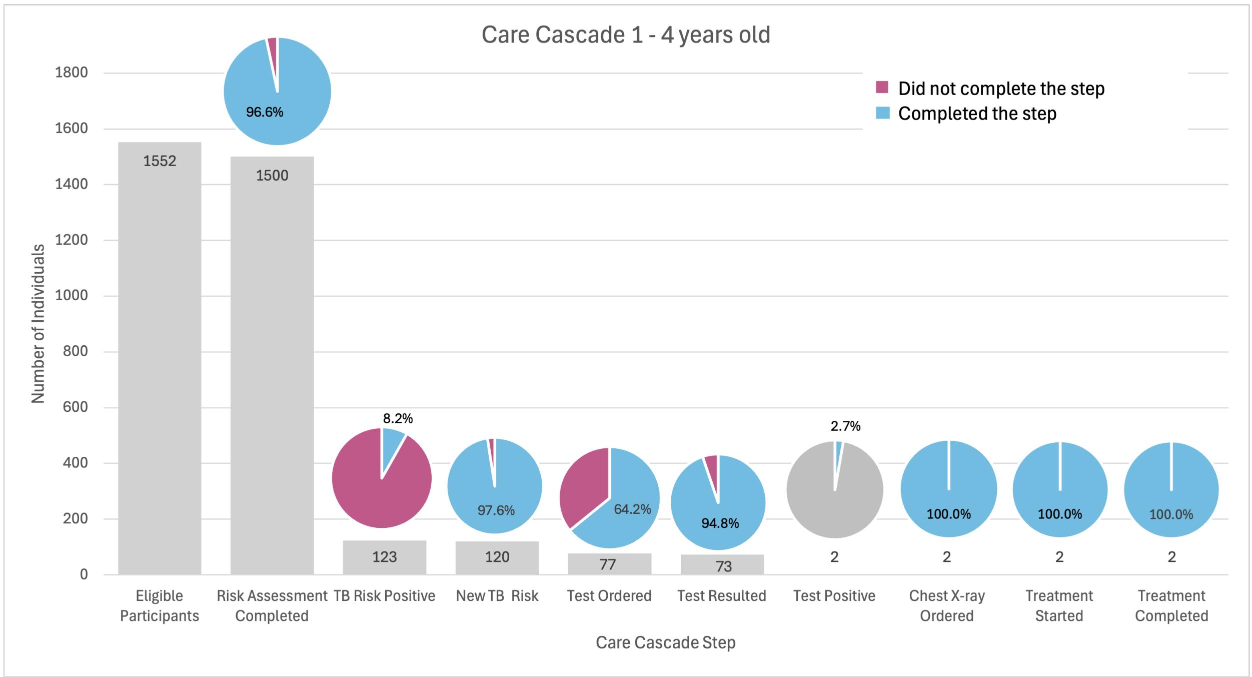


**B.**

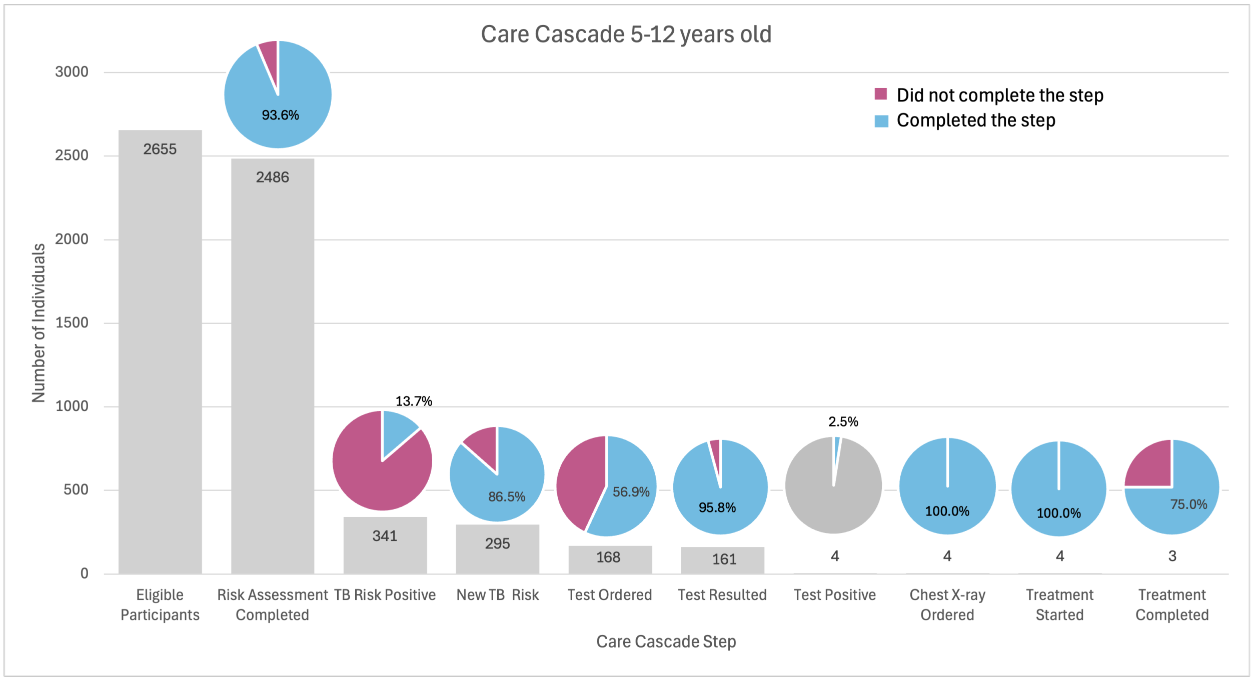


**C.**

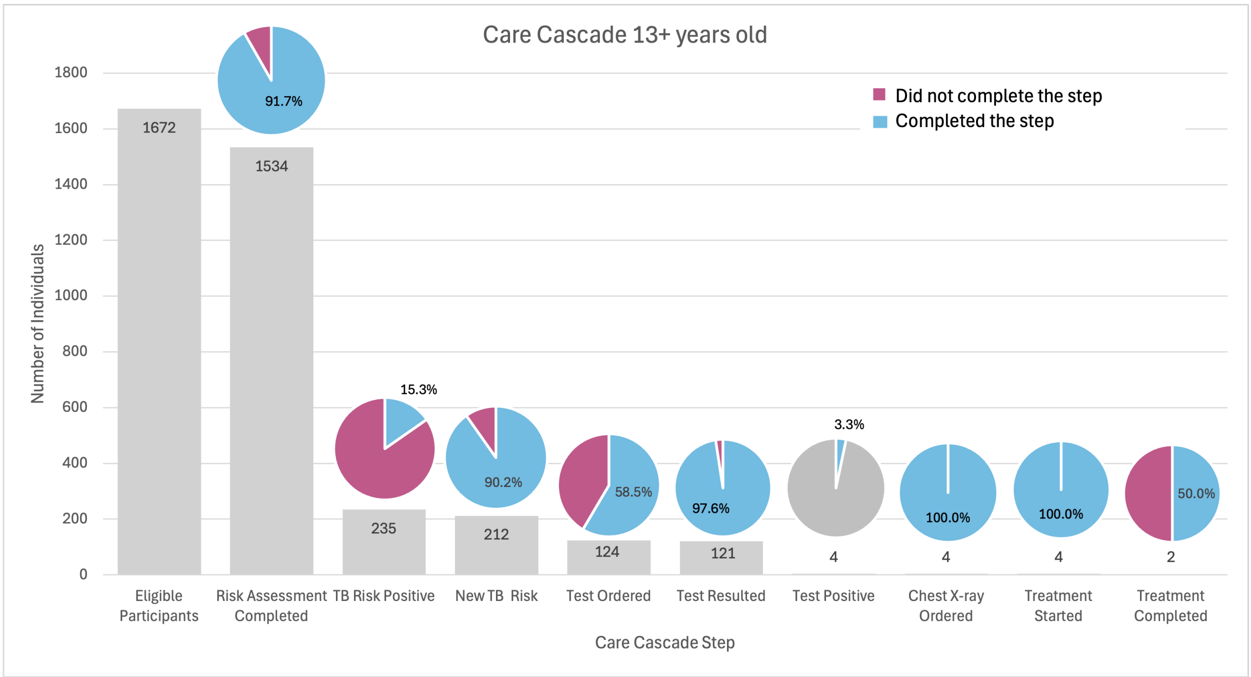
Care Cascade 13+ years old
Did not complete the step
Completed the step
Number of Individuals
0
200
400
600
800
1000
1200
1400
1600
1800
1672
1534
235
212
124
121
4
4
4
2
91.7%
15.3%
90.2%
58.5%
97.6%
3.3%
100.0%
100.0%
50.0%
Eligible Participants
Risk Assessment Completed
TB Risk Positive
New TB Risk
Test Ordered
Test Resulted
Test Positive
Chest X-ray Ordered
Treatment Started
Treatment Completed
Care Cascade Step

**Supplemental Figure 2.** Care Cascade for pediatric LTBI by primary language: A) English (n = 4,253); or B) Non English (n=1,296)

**A.**

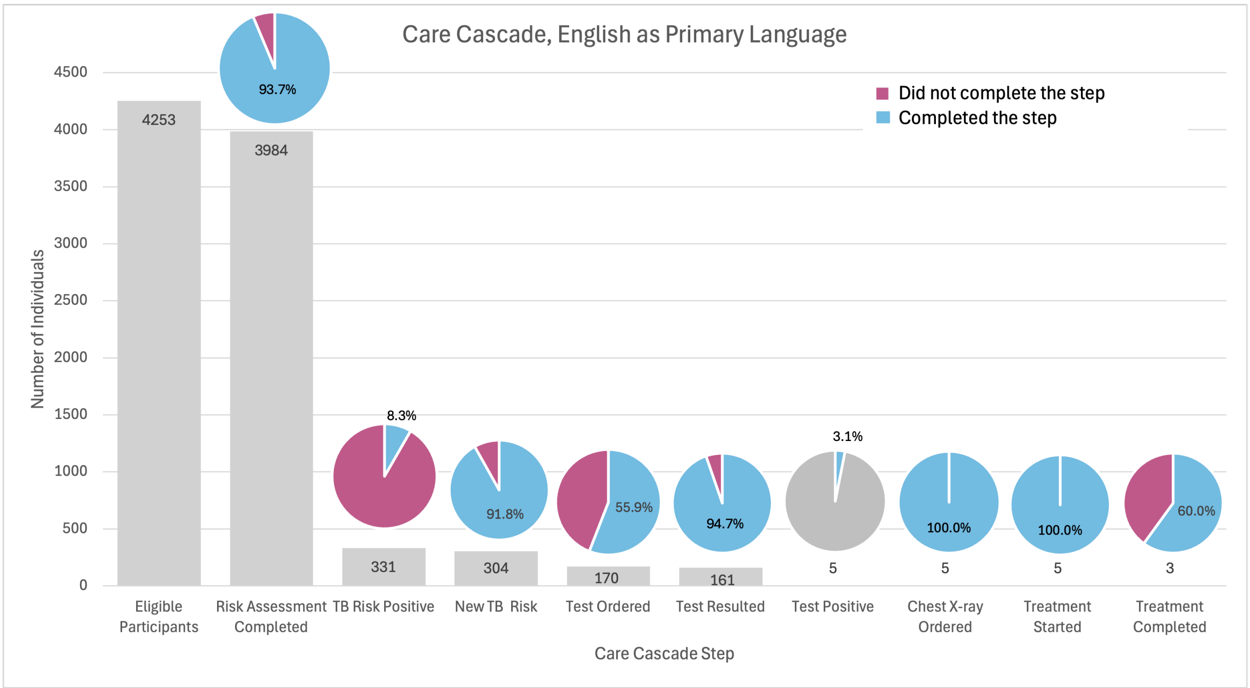


**B.**

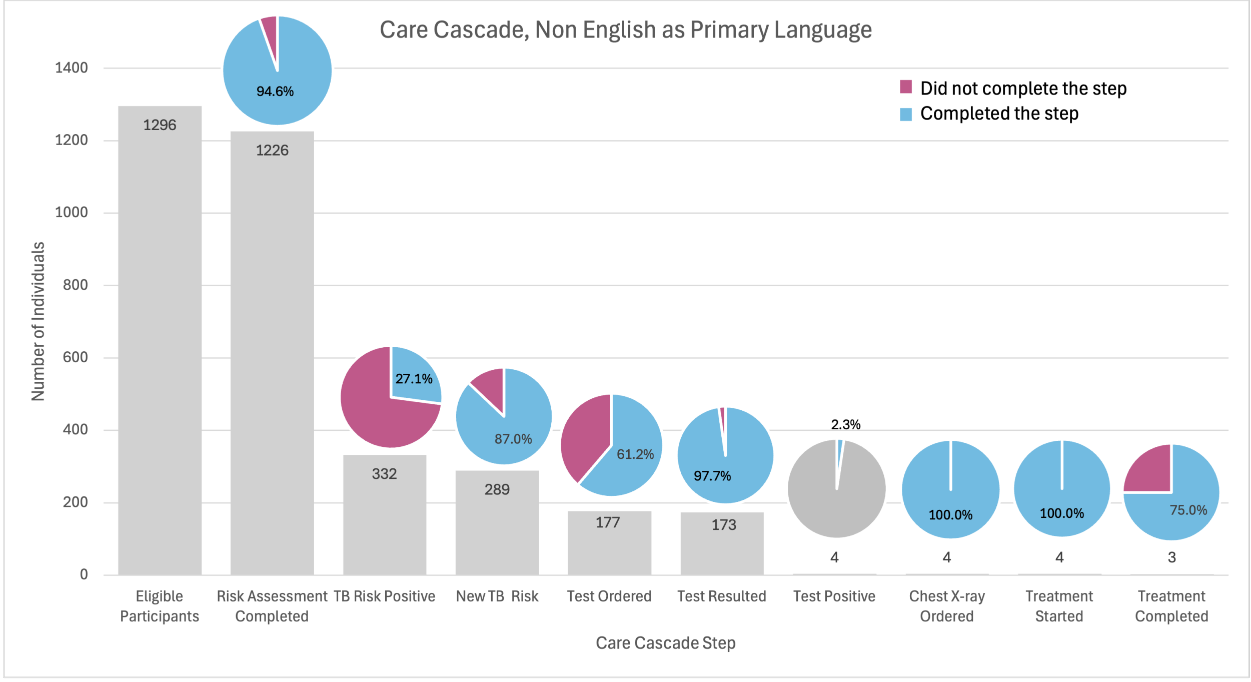
